# Plasma Metabolite Associations with Incident Heart Failure with Reduced and Preserved Ejection Fraction

**DOI:** 10.64898/2026.07.20.26358530

**Authors:** Bhavya Chebrolu, Priyam Choksi, Salvatore Carbone, Brittany N. Weber, Jessica Lasky-Su, Elizabeth W. Karlson, Sheila Hegde, Leo F. Buckley

**Author notes:** <u>Address for Correspondence</u>: Leo F. Buckley, PharmD MPH, Translational Cardiovascular Epidemiology Research Unit, Division of Cardiology, University of Texas Southwestern Medical Center, Department of Epidemiology, UT Southwestern Peter O’Donnell Jr. School of Public Health Dallas, TX 75390. <u>Disclosures</u>: BNW is supported by NIH/NHLBI K23HL159276, AHA CDA851511 and the AHA Recurrent Pericarditis Initiative and reports consulting or advisory fees from Novo Nordisk, Bristol Myers Squibb and Horizon Therapeutics. JL-S is a scientific advisor for Precion, Inc and a consultant to Tru Diagnostic, Inc. SMH has received advisory board fees from Cytokinetics and her institution has received fees for core laboratory services from Cytokinetics and Bristol Myers Squibb. LFB is supported by NIH/NHLBI K23HL150311 and reports consulting fees from scPharmaceuticals and continuing education fees from ASHP Advantage. The remaining authors have nothing to disclose.

## Abstract

**Background:** Prior studies on metabolite associations with incident heart failure (HF) used billing code-based definitions and lacked the data on left ventricular ejection fraction needed to determine associations with HF with reduced (HFrEF) and preserved (HFpEF) ejection fraction.

**Objectives:** Identify potentially causal plasma metabolite associations with HFrEF and HFpEF, ascertained using a validated machine learning- and natural language processing-based algorithm, in up to 38,000 individuals.

**Methods:** We included MGB Biobank participants who had available metabolomics data and no history of HF at baseline. The primary exposures were plasma levels of 42 metabolites measured using a H^1^ nuclear magnetic resonance platform. The primary outcome was incident HF, ascertained by a validated machine learning- and natural language processing-based algorithm. Multivariable Cox proportional hazards regression to quantify the associations between a 1-SD difference in metabolite level and the time to incident HF. Mendelian randomization analysis was used test the potential causality of each metabolite-HF association.

**Results:** The final analytical cohort included 38,628 individuals with a mean age of 63 years (56% women). Higher plasma levels of glutamine associated with a higher risk of incident HF (HR [95% CI]: 1.21 [1.07-1.35]) while higher levels of docosahexaenoic acid (an omega-3 fatty acid) (0.85 [0.75-0.95]), phosphatidylcholines (0.85 [0.75-0.97]), phosphoglycerides (0.86 [0.76- 0.97]) and total cholines (0.85 [0.75-0.97]) associated with a lower HF risk. Docosahexaenoic acid and total omega-3 fatty acids associated with HFpEF. Associations with HFpEF tended to be stronger than with HFrEF for several fatty acids, including omega-3 fatty acids, in individuals with obesity, but not coronary artery disease or diabetes. Mendelian randomization analysis supported causal associations between higher levels of docosahexaenoic acid and omega-3 fatty acids with a higher risk of HF and greater left ventricular mass index.

**Conclusions:** Dysregulated omega-3 fatty acid metabolism may be causally associated with a higher risk of incident HFpEF.

**Condensed Abstract:** We investigated metabolic contributors to incident heart failure (HF) in the MGB Biobank, leveraging metabolomics and a validated machine learning/NLP algorithm for HF ascertainment. Among 38,628 participants without HF at baseline, plasma levels of 42 metabolites were evaluated using multivariable Cox models and Mendelian randomization. Higher glutamine levels were associated with increased HF risk, while docosahexaenoic acid (DHA), phosphatidylcholines, phosphoglycerides, and total cholines were inversely associated. DHA and total omega-3 fatty acids were linked to HFpEF. Mendelian randomization suggested causal associations between higher DHA and omega-3 fatty acid levels and elevated HF risk, implicating dysregulated omega-3 metabolism in HF development.

## Introduction

Dysregulated metabolism contributes to the pathogenesis of heart failure.^1^ Treatment with drugs that modulate cardiac metabolism, such as SGLT2 inhibitors and GLP-1R agonists, can prevent the development of heart failure and its progression.^2^ The growing prevalence of cardiometabolic risk factors creates a need for a deeper understanding of the metabolic pathways that promote cardiac dysfunction and heart failure, which may inform the development of biomarkers and therapeutic targets.^3^

Unbiased analyses of dozens and hundreds of circulating metabolites (metabolomics) in observational cohort studies can accelerate research on metabolism and heart failure.^4–7^ However, more rigorous studies on this topic are needed as prior studies used small sample sizes, ascertained heart failure events with medical billing codes and were unable to distinguish between heart failure with reduced (HFrEF), mid-range or preserved ejection fraction (HFpEF).^4–7^ Moreover, the observational cohort designs of prior studies may be affected by residual confounding.

The Mass General Brigham (MGB) Biobank has enrolled over 160,000 individuals from across southeastern New England region. The MGB Biobank has metabolomics data available for approximately 48,000 participants, a validated machine learning- and natural language processing-based algorithm for heart failure case ascertainment and linked electronic health record data for categorization of HFrEF and HFpEF. Mendelian randomization analysis can support causal inference on the effects of metabolites levels on heart failure risk. We conducted a metabolomics analysis in the MGB Biobank and performed Mendelian randomization analysis to identify potentially causal metabolic contributors to the development of heart failure, including HFrEF and HFpEF.

## Methods

### Study Cohort

We performed an observational cohort study of the participants in the MGB Biobank. MGB Biobank participants were included in the analysis if they had plasma metabolomics data available and a body mass index between 12 and 78.4 kg/m^2^. Participants were excluded if they had a history of heart failure, were taking omega-3 fatty acids or hormonal contraceptives in the prior 3 months or were pregnant at the time of blood draw. This study was deemed exempt by the MGB Human Subjects Research Board.

### Exposures

Metabolite levels were quantified using a targeted commercial H1 nuclear magnetic resonance (NMR) platform (Nightingale Health, Helsinki, Finland).^8^ Of the 249 metabolites and metabolite ratios reported from this platform, we excluded metabolite ratios and atherogenic lipids, cholesterols and lipoproteins to identify coronary artery disease-independent metabolite- heart failure associations. The primary exposures of interest were plasma levels of 42 metabolites (**Supplemental Table 1**).

Analysis of raw biomarker distributions across batches and spectrometers suggested minimal batch- and spectrometer-related effects (**Supplemental Figure 1**). The levels of clinically available metabolites measured by the NMR platform correlated significantly with levels available in the electronic health record (**Supplemental Figure 2**).

### Outcomes

The primary outcome of interest was incident heart failure overall (i.e., any left ventricular ejection fraction). Heart failure cases were identified using a validated machine learning- and natural language processing-based algorithm that was trained on the electronic health records of 548 confirmed cases of heart failure as previously described.^9^ In brief, an automated feature extraction protocol was used to identify 1,257 candidate heart failure-related concepts, such as comorbidities, symptoms and medications, from publicly available databases. Of these 1,257 concepts, 16 were frequent enough in the electronic health records of the 548 confirmed heart failure cases to be used in model training. The model also considered International Classification of Diseases (ICD) codes and prescription records. A logistic regression classifier with adaptive least absolute shrinkage and selection operator (LASSO) was used to build a model for the predicted probability of a positive history of heart failure.

The final heart failure model was based on the number of the heart failure-related ICD codes and the number of mentions of heart failure-related Unified Medical Language System terms (“heart failure”: C0018802; “ischemic cardiomyopathy”: C0349782; and “loop diuretics”: C0354100”) with an additional term for the total number of hospital encounters with an ICD code to account for the interindividual differences in the density of electronic health record data.^9^ For the present analysis, individuals were considered to have heart failure if their predicted probability of heart failure was 0.90 or greater. This implementation of the algorithm had a positive predictive value of 0.90, a sensitivity of 0.53 and an area-under-the-curve (AUC) of 0.921 in the 548-case training dataset. In comparison, use of one or more heart failure-related ICD codes had a positive predictive value of 0.23.

Heart failure cases were categorized as heart failure with reduced ejection fraction (HFrEF; left ventricular ejection fraction less than 40%) or heart failure with mid-range or preserved ejection fraction (HFpEF; left ventricular ejection fraction of 40% or greater). The left ventricular ejection fraction value was extracted from the echocardiogram report that was closest in time to the heart failure diagnosis but no greater than 90 days before or after the diagnosis date. Left ventricular ejection fraction data were available for 196 of the total 375 incident heart failure events (47 HFrEF; 149 HFpEF). Individuals with uncategorized heart failure were assumed to be heart failure-free in these analyses.

### Covariates

Demographic covariates were derived from patient-reported date of birth, patient-selected sex at birth (male, female or X), patient-selected race (American Indian or Alaska Native, Asian, Black, Native Hawaiian or other Pacific Islander, White and Other, Unknown or Declined) and patient-selected ethnicity (Hispanic, Non-Hispanic and Unknown or Declined).

Medical history was abstracted from ICD codes in the electronic health record data. The presence of one or more ICD codes in the 12 months before the date of the metabolomics blood draw was considered to indicate a positive history for that condition. Medication history was abstracted from electronic health record prescription orders. MGB Biobank participants were considered to be taking a medication if they had one or more prescription orders for that medication in the 90 days before the date of metabolomics blood draw. The lone exception to the medication use definition was for steroids, which required at least two prescription orders to capture chronic, rather than one-time, users.

The years in which the blood sample used for metabolomics analysis was drawn were grouped into quartiles to account for temporal confounding. Fasting status was collected at the time of blood draw for all participants.

### Statistical Analysis

Participant characteristics were described using numbers and percentages or means and standard deviations (SD). Prior to analysis, metabolite measurements of 0 or “NA” were imputed with half of the minimum value for that metabolite. Outliers more than 4-times the interquartile range from the median were excluded. The metabolite measurements were then natural log-transformed and a linear regression model was used to remove spectrometer effects. The model residuals were scaled and used in downstream analysis.

We used Cox proportional hazards regression to quantify the associations between a 1- SD difference in metabolite level and the time to incident heart failure, incident HFrEF and incident HFpEF. We constructed models that included age, sex, race, ethnicity, plasma creatinine, body mass index, year of blood draw quartile, fasting status, total number of hospital encounters with an ICD code, statin use, coronary artery disease, diabetes, dyslipidemia, hypertension, atrial fibrillation and current use of an angiotensin-converting enzyme or angiotensin receptor blocker, thiazide diuretic, calcium channel blocker, beta blocker, mineralocorticoid receptor antagonist or disease-modifying anti-rheumatic drug.

We assessed effect modification by diabetes status, coronary artery disease history and obesity status (<30 kg/m^2^ vs. ≥30 kg/m^2^) using a multiplicative interaction term in the Cox proportional hazards regression models. All analyses used the Benjamini-Hochberg procedure to adjust for multiple testing with a q-value < 0.05 considered statistically significant.

### Mendelian Randomization Analysis

We performed Mendelian randomization analysis to test the potential causality of metabolite-heart failure associations observed in the MGB Biobank (adjusted P<0.10). We used summary statistics from a genome-wide association study of circulating metabolite levels^10^ to build genetic instruments for each heart failure-related metabolite using metabolite quantitative trait loci (mQTL). mQTLs within a 10 kb window with linkage disequilibrium r^2^ calculated using the European superpopulation within the 1000 Genomes Project of 0.001 or greater were clumped and the pQTL with the smallest P-value was retained. Heart failure summary statistics were derived from a meta-analysis of heart failure genome-wide association studies.^11^ Cardiac structure and function summary statistics were obtained from genome-wide analyses of the UK Biobank Imaging Study.^12–14^

The primary Mendelian randomization estimator was derived from inverse variance weighted meta-analysis of mQTL-outcome associations. We used the weighted median estimator and the weighted mode estimator to derive estimates robust against up to 50% invalid instruments. Horizontal pleiotropy was detected as a statistically significant intercept from MR Egger regression. For the Mendelian randomization analysis, metabolite associations with heart failure or cardiac structure and function measures were considered statistically significant and robust if the inverse variance weighted meta-analysis was statistically significant (P<0.05), at least one of the weighted median or weighted mode estimators was statistically significant (P<0.05) and the MR Egger intercept was not statistically significant (P>0.05).

## Results

### MGB Biobank Participant Characteristics

The final analytical cohort included 38,628 individuals (**Supplemental Figure 3**). The mean (SD) age was 63 (16) years, 56% were women and 87% were White. Hypertension (65%), dyslipidemia (55%), diabetes (31%) and coronary artery disease (15%) were the most common cardiometabolic comorbidities.

### Metabolite Associations with Incident Heart Failure in the MGB Biobank

A total of 375 incident heart failure events occurred over a median follow-up of 6 (5-9) years (1.5 [1.4-1.7] events per 1,000 person-years). There were 5 statistically significant metabolite associations with incident heart failure after multiple testing correction (**Supplemental Figure 4**). Higher plasma levels of glutamine (HR [95% CI]: 1.21 [1.07-1.35]) associated with a higher risk of incident heart failure while lower levels of docosahexaenoic acid (an omega-3 fatty acid) (HR [95% CI]: 0.85 [0.75-0.95]), phosphatidylcholines (HR [95% CI]: 0.75-0.97), phosphoglycerides (HR [95% CI]: 0.86 [0.76-0.97]) and total cholines (HR [95% CI]: 0.85 [0.75- 0.97]) associated with a higher heart failure risk (**Figure 1**).

**Figure 1.**
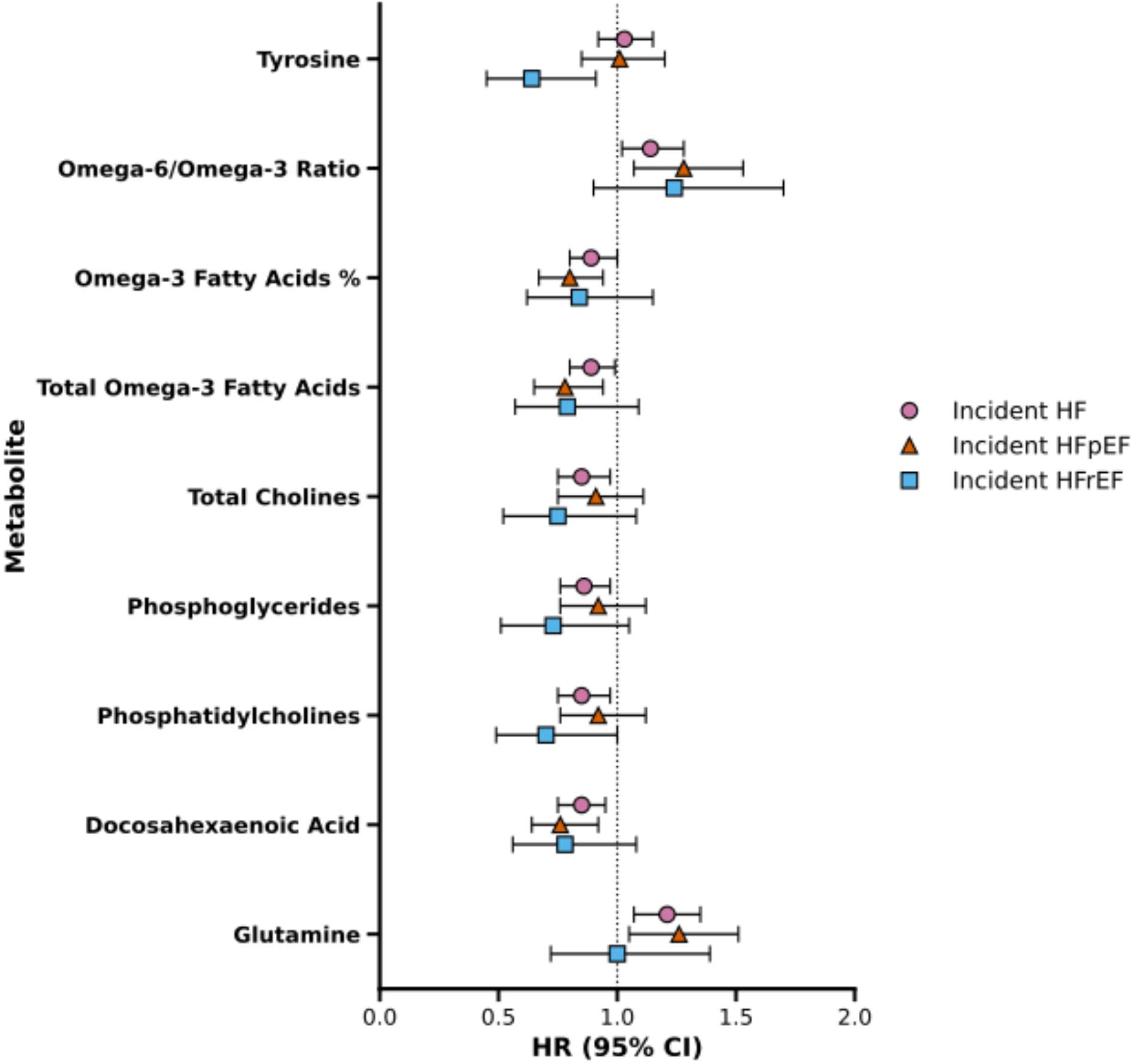
Metabolite Associations with Incident Heart Failure. This forest plot summarizes the significant associations between plasma metabolites and incident HF, incident HFrEF and incident HFpEF. The confidence intervals have not been corrected for multiple testing. CI = confidence interval; HF = heart failure; HFpEF = HF with preserved ejection fraction; HR = hazard ratio; HFrEF = heart failure with reduced ejection fraction

### Metabolite Associations with Incident Heart Failure with Reduced and Preserved Ejection Fraction in the MGB Biobank

There were 149 incident HFpEF events (0.6 [0.5-0.7 events per 1,000 person-years] and 45 incident HFrEF events (0.2 [0.1-0.2] events per 1,000 person-years). Docosahexaenoic acid, glutamine, total omega-3 fatty acids, percentage of omega-3 fatty acids and the ratio of omega- 6 to omega-3 fatty acids significantly associated with incident HFpEF (**Figure 1**). Tyrosine was the only metabolite that significantly associated with HFrEF (**Figure 1**).

### Effect Modification of Metabolite Associations with Incident Heart Failure in the MGB Biobank

Neither diabetes status nor coronary artery disease status significantly modified any of the associations between plasma metabolites and incident heart failure overall, incident HFrEF or incident HFpEF (**Supplemental Table 3** and **Supplemental Table 4**).

Higher alanine levels were associated with a higher risk of incident heart failure overall in participants without obesity (HR [95% CI]: 1.22 [1.04-1.44]) but a lower risk in participants with obesity (HR [95% CI]: 0.84 [0.73-0.97]; P-interaction = 0.021). Higher saturated fatty acids as a percentage of total fatty acids was associated with a greater risk of incident HFrEF in non-obesity (HR [95% CI]: 2.07 [1.21-3.53]) but not obesity (HR [95% CI]: 0.74 [0.53-1.02]; P-interaction = 0.035). Higher levels of monounsaturated fatty acids, omega-3 fatty acids, phosphatidylcholines, phosphoglycerides, saturated fatty acids and total fatty acids associated with a lower risk of incident HFpEF in participants with obesity only (**Supplemental Table 5**), but the interaction was not statistically significant (P-interaction < 0.10 for each).

### Causal Metabolite Associations with Heart Failure in Mendelian Randomization Analysis

Mendelian randomization analysis was performed for the 5 metabolites that significantly associated with incident heart failure (docosahexaenoic acid, glutamine, phosphatidylcholines, phosphoglycerides and total cholines) plus 2 metabolites with P > 0.05 and < 0.10 (total omega- 3 fatty acids and phenylalanine; no available instrumental variables for the ratio of omega- 6/omega-3 fatty acids). Mendelian randomization analysis supported causal associations between higher levels of docosahexaenoic acid and omega-3 fatty acids with a higher risk of heart failure (**Figure 2**). Higher omega-3 fatty acid levels were associated with greater left ventricular mass index in Mendelian randomization analysis (**Figure 2**). **Supplemental Table 6** summarizes the Mendelian randomization analysis results for all metabolites.

**Figure 2.**
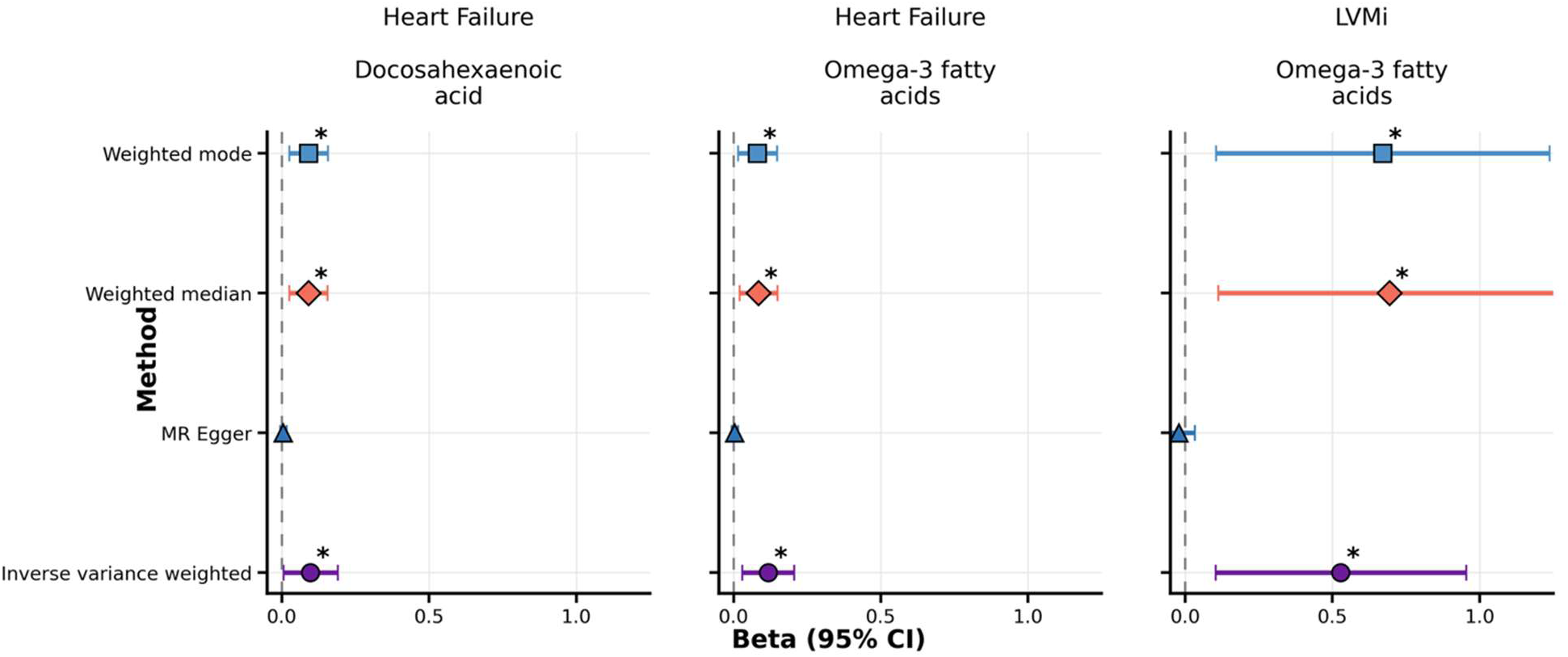
Mendelian Randomization Results. This Forest plot summarizes the estimated causal association between various metabolites and the risk of heart failure or with measures of cardiac structure and function. The plot only shows results where the metabolite-outcome associations were significant in inverse variance-weighted meta-analysis and either weighted median or weighted mode MR analyses and where the MR Egger intercept was non-significant. CI = confidence interval; MR = Mendelian randomization

## Discussion

We leveraged biobank-scale plasma metabolomics data and a validated machine learning- and natural language processing-based heart failure algorithm in 38,628 MGB Biobank participants to identify potentially causal metabolite associations with incident heart failure, including sub-phenotyping for HFrEF and HFpEF. Our analysis supports a causal role for omega-3 fatty acids, in particular docosahexaenoic acid, in the pathogenesis of heart failure and left ventricular hypertrophy. In addition, our results provide stronger evidence for metabolite associations with HFpEF than HFrEF and suggest possible effect modification by obesity status. By identifying metabolites potentially causally related to the pathogenesis of heart failure, our study has opened new avenues of research for therapeutic targets and biomarkers.

Our findings build upon the existing literature by providing causal evidence for an effect of omega-3 fatty acids, in particular docosahexaenoic acid, on the risk of incident heart failure and by investigating HFrEF and HFpEF separately. While the Gruppo Italiano per lo Studio della Sopravvivenza nell’Infarto Miocardico-Heart Failure (GISSI-HF) trial demonstrated a 9% reduction in the risk of all-cause death and an 8% reduction in the risk of all-cause death or cardiovascular hospitalization with omega-3 fatty acid supplementation (850 mg eicosapentaenoic acid and 882 mg docosahexaenoic acid), the study population already had established heart failure.^15^ In the VITamin D and OmegA-3 TriaL (VITAL), which enrolled an unselected general population, omega-3 fatty acid supplementation (380 mg docosahexaenoic acid and 460 mg eicosapentaenoic acid) decreased the risk of incident heart failure in patients with, but not without, type 2 diabetes at baseline.^16,17^ Higher omega-3 fatty acid doses may have enhanced the potential benefits observed in VITAL.^18^ An analysis of the Multi-Ethnic Study of Atherosclerosis (MESA) found that higher eicosapentaenoic acid levels, another omega-3 fatty acid, associated with a lower risk of incident heart failure.^19^ Our analysis strengthens the available evidence by using Mendelian randomization analysis for causal inference and analyzing HFrEF and HFpEF separately. Omega-3 fatty acid supplementation has further appeal as a primary prevention strategy due to the potential for non-pharmacologic interventions, such as dietary modifications, that would appeal to a broad primary prevention population.^20^ Interestingly, higher levels of monounsaturated fatty acids and omega-3 fatty acids associated with a lower risk of HFpEF only in the subgroup of participants with obesity. Prior clinical and pre-clinical research has implicated unsaturated fatty acids in the pathogenesis of obesity-related HFpEF.^21,22^ Given the growing burden of obesity and HFpEF, non-pharmacological dietary interventions^20^ to modulate unsaturated fatty acid intake may have potential to improve cardiac dysfunction and prevent HFpEF. Our findings provide strong rationale for randomized controlled trials testing high-dose omega-3 fatty acid supplementation for the primary prevention of heart failure in the general population and targeted dietary interventions in individuals with obesity.

Whereas dysregulated metabolism potentially contributes directly to HFpEF pathogenesis, HFrEF may result more often from the ischemic consequences of metabolic risk factors such as diabetes. Thus, our analysis extends the existing literature in an important direction by investigating metabolite associations with HFpEF and HFrEF. Several prior studies on this topic did not distinguish between HFpEF and HFrEF.^4–6^ One study used an untargeted metabolomics platform to identify 21 metabolite-HFpEF associations in the Atherosclerosis Risk in Communities (ARIC) prospective cohort study with 117 physician-adjudicated HFpEF events and 101 physician-adjudicated HFrEF events.^7^ Our study, in contrast, used a targeted metabolomics platform and derived HFpEF (n=149) and HFrEF (n=45) outcomes from clinically available electronic health records and echocardiography reports. While our cohort may have broader generalizability to routine clinical practice, the ARIC metabolomics study had a wider breadth of metabolites. We also extend the literature by examining obesity as an effect modifier of the metabolite-HFpEF and metabolite-HFrEF associations, an important analysis given the public health relevance of obesity and HFpEF.

Our study has distinct strengths and limitations. We used a large sample size, an established metabolomics platform, a validated heart failure case ascertainment algorithm and causal inference methodology. Broader racial and ethnic diversity and enrollment of patients receiving care outside of the tertiary academic medical center setting would enhance the generalizability of our results. A higher proportion of participants with echocardiography data would have increased our power in the HFrEF and HFpEF analyses. Our analysis re-coded individuals with heart failure who were missing echocardiography results as not having developed heart failure, which may have biased our results towards the null.

## Conclusions

Biobank-scale plasma metabolomics data, a validated machine learning- and natural language processing-based heart failure algorithm and Mendelian randomization analysis provide robust evidence to support a causal role of omega-3 fatty acids, in particular docosahexaenoic acid, in the pathogenesis of incident heart failure. These metabolites may have a role as biomarkers of risk prediction or treatment response and counteracting dysregulation in these pathways may prevent the development of incident heart failure.

## Data Availability

Access to the MGB Biobank is limited to MGB investigators and approved collaborators.

---

**Table 1.** Mass General Brigham Biobank Participant Characteristics.

| **Characteristic** | **N (%) or Mean (Standard Deviation)** |
| --- | --- |
| **Age, years** | 63 (16) |
| **Sex** |  |
| Female | 21,436 (56) |
| Male | 17,184 (44) |
| X | 2 (<1) |
| **Self-Selected Race** |  |
| American Indian or Alaska Native | 96 (<1) |
| Asian | 951 (2) |
| Black | 1,812 (5) |
| Native Hawaiian or Other Pacific Islander | 14 (<1) |
| Other, Unknown or Declined | 2,302 (6) |
| White | 33,447 (87) |
| **Self-Selected Ethnicity** |  |
| Hispanic | 573 (1) |
| Non-Hispanic | 34,427 (89) |
| Unknown or Declined | 3,622 (9) |
| **Medical History** |  |
| Coronary Artery Disease | 5,937 (15) |
| Diabetes | 12,005 (31) |
| Dyslipidemia | 21,310 (55) |
| Hypertension | 25,301 (65) |
| Stroke or Transient Ischemic Attack | 1,403 (4) |
| Acute Myocardial Infarction | 602 (2) |
| Rheumatoid Arthritis | 1,882 (5) |
| Psoriasis | 1,281 (3) |
| Chronic Kidney Disease | 2,526 (7) |
| Atrial Fibrillation | 2,714 (7) |
| **Current Medications** |  |
| Angiotensin Receptor Blocker | 5,181 (13) |
| Angiotensin-Converting Enzyme Inhibitor | 3,433 (9) |
| Mineralocorticoid Receptor Antagonist | 338 (1) |
| Thiazide Diuretic | 2,214 (6) |
| Calcium Channel Blocker | 2,382 (6) |
| Beta Blocker | 4,031 (10) |
| Steroids | 2,895 (7) |
| Non-Steroidal Disease-Modifying Anti-Rheumatic Drug | 2,649 (7) |
| Statin | 6,230 (16) |

